# Machine learning methods to track dynamic facial function in facial palsy

**DOI:** 10.1101/2024.03.16.24304357

**Authors:** Akshita A. Rao, Jacqueline J. Greene, Todd P. Coleman

## Abstract

For patients with facial paralysis, the wait for return of facial function and resulting vision risk from poor eye closure, difficulty speaking and eating from flaccid oral sphincter muscles, as well as psychological morbidity from the inability to smile or express emotions through facial movement can be devastating. There are limited methods to assess ongoing facial nerve regeneration: clinicians rely on subjective descriptions, imprecise scales, and static photographs to evaluate facial functional recovery, and thus facial nerve regeneration remains poorly understood. We propose a more precise evaluation of dynamic facial function through video-based machine learning analysis to facilitate a better understanding of the sometimes subtle onset of facial nerve recovery and improve guidance for facial reanimation surgery.

**Methods:** We here present machine learning methods employing likelihood ratio tests, optimal transport theory, and Mahalanobis distances to: 1) assess the use of defined facial landmarks for binary classification of different facial palsy types; identify regions of asymmetry and potential paralysis during specific facial cues; and 3) quantify palsy severity and map it directly to widely used clinical scores, offering clinicians an objective way to assess facial nerve function.

**Results:** Our work presents promising results of utilizing videos, rather than static photographs, to provide robust quantitative analyses of dynamic properties for various facial movements without requiring manual assessment.

**Significance:** The long-term potential of this project is to enable clinicians to have more accurate and timely information to make decisions for facial reanimation surgery which will have drastic consequences on the quality of life for affected patients.

## I. Introduction

There are an estimated over 500,000 cases of peripheral nerve transection requiring repair annually in the United States [1] and peripheral nerve injuries cost the US healthcare system $150 billion annually [2]. Despite this, the current understanding of the spectrum of nerve injury and regenerative outcomes is based on clinical observational data alone, with unfortunately little objective data to guide surgical intervention [3]. Acquired facial paralysis occurs in over 150,000 Americans every year from a variety of causes: Bell’s Palsy, benign or malignant tumors, trauma, infections and neurologic and iatrogenic injuries [4].

While facial nerve (FN) recovery is robust in Bell’s Palsy and should be managed conservatively, for many other conditions, the prognosis is much more uncertain and may take months to years to complete. In such cases, a critical time window for successful facial reanimation surgery is passing due to muscle degeneration and atrophy [5]. As detecting the onset of nerve regeneration and muscle reinnervation can be subtle and difficult to assess clinically, determining prognosis for FN recovery represents a black-box scenario with limited data to guide clinicians and patients. In the setting of no overt visible facial movement, the choice to proceed with facial reanimation surgery, such as a cranial nerve V-VII transfer, is fraught with difficulty [6] [7] [8] [9].

There are limited objective methods to quantify FN function; clinicians rely on subjective descriptions, imprecise scales, and static photographs to evaluate facial functional recovery [10] [11] [12] [13] [14]. There are no current imaging modalities to detect the onset, progression, or failure of FN regeneration [3].

The lack of objective FN outcomes data and lengthy recovery times has hampered the field of facial reanimation surgery for some time [15]. For example, only 24% of patients requiring FN sacrifice during parotid tumor excision receive FN repair [16]. For patients with facial paralysis, the wait for return of facial function and the resulting vision risk from poor eye closure, difficulty speaking and eating from flaccid oral sphincter muscles, as well as the psychological morbidity from the inability to smile or express emotions through facial movement, can be devastating. Experts in the field indicate that a reliable objective system could truly revolutionize the way clinicians diagnose and monitor patients suffering from such FN injuries [17].

### A. Applications of Machine Learning in Analysis of Facial Palsy

Machine learning (ML) algorithms for facial analysis have advanced significantly in the past few decades [18] [19], but most are based on training datasets of intact facial function. When applied to photographs of patients with facial paralysis, these tools have been shown to cause significant landmark inaccuracy, precluding clinical utility [20] [21]. The most advanced method of automated facial analysis in facial paralysis patients, Emotrics, is limited to static photos only and requires significant manual landmark adjustments [21]. Other attempts to quantify facial function have incorporated proprietary marketing software to estimate facial emotional expressions [22] [23] [24] [25] or were limited to a single surgical case report [26] following surgical interventions.

While there has been significant progress in using video-based ML approaches to assess facial asymmetry in stroke survivors [27], [28], these studies focus primarily on neuromuscular disorders rather than facial palsy. In the context of facial palsy, an objective, open-source, and rigorous quantification of dynamic facial function in videos remains elusive [26] [30]. This gap persists due to challenges in accumulating sufficiently large and diverse training datasets in the field.

There is an unmet need to develop a more precise evaluation of dynamic facial function to facilitate a better understanding of the spectrum of facial palsy, improve guidance for and outcomes following facial reanimation surgery, and detect the subtle onset of FN recovery, for which there currently is no reliable test [15]. Contrary to contemporary beliefs regarding the limitations of ML algorithms, recent open-source computer vision and ML algorithms available through Python (OpenCV, dlib) [19] [31] applied to facial palsy patients revealed surprisingly robust landmark accuracy despite significant facial asymmetry [32]. Specifically, a recent finding demonstrated a significant reduction in landmark error rate in videos compared to photos in a standardized data set across all facial palsy severity types, in part because the volume of data available from a video of a standard FN exam is orders of magnitude larger than static photography (from 8 photos to 3600 frames for a 60 second video recorded at 60fps) [32]. However, continuous video data also presents challenges. Unlike static images captured at peak facial expressions, where landmark localization is inherently more precise, videos often include frames where facial expressions are incomplete or transitioning, leading to variability in landmark accuracy [33]. Finetuning facial landmark models has been shown to improve performance on both static images and videos, but videos remain particularly advantageous for detecting subtle, dynamic changes in facial function over time [33]. These trade-offs underscore the need for methods that leverage the strengths of both approaches to enable precise, clinically meaningful analysis of facial palsy.

Building upon recent accomplishments in reducing landmark error rates in videos of patients with facial palsy, this study introduces a comprehensive framework that bridges statistical methods, machine learning, and clinical applicability to analyze and quantify dynamic facial function. Specifically, we present novel methodologies that include likelihood ratio tests, optimal transport theory, and Mahalanobis distances to objectively measure and classify facial function, making substantial strides in addressing the current gaps in facial palsy assessment. Our methods translate ML-based analyses into clinically actionable insights, potentially aiding in facilitating decisions during facial reanimation surgeries. We provide patient-specific dynamic facial function evaluation and demonstrate that our methods show improved performance when tailored to specific contexts in the clinically relevant facial nerve exam. To further demonstrate clinical relevance, we validate our methods on surgical case studies and demonstrate utility in predicting widely used clinical scores. Our pipeline provides a foundation for automated and scalable video-based analysis, reducing reliance on manual annotations and subjective scoring. This positions the tool as a future-ready option for clinics aiming to incorporate precision-based evaluation systems. To summarize, the main contributions of this paper include:

- We demonstrate, for the first time, the use of Wasserstein barycenters to quantify the severity of facial asymmetry and map it directly to clinical scores, offering clinicians a robust and objective way to assess FN recovery.
- Using machine learning, we achieve binary classification of different facial palsy types with high accuracy, uniquely distinguishing not only between healthy controls and facial palsy but also between different palsy types (i.e. flaccid and non-flaccid/synkinesis)— a task not previously accomplished with this level of accuracy.
- Beyond theoretical analyses, we validate our methods on two surgical case studies, demonstrating utility in predicting clinical scores (e.g., House-Brackmann, Sunnybrook, and eFACE) and assessing surgical outcomes in a longitudinal manner.

In **Methods**, we provide details on the selection of facial landmarks, statistical and ML methods to analyze these landmarks, and our novel use of Wasserstein barycenters for dynamic facial function analysis and clinical score prediction. In **Results**, we demonstrate how the presented methods allow us to classify between different facial palsy types, map palsy types to clinical scores with high reliability, and apply these analyses to individual surgical case studies. Finally, in **Discussion**, we explain the implications and limitations of our work and suggest avenues for future directions.

## II. Methods

### A. Data Collection

Videos of healthy controls and facial palsy patients clinically diagnosed with facial palsy were prospectively gathered. The Massachusetts Eye and Ear Infirmary (MEEI) Facial Palsy Photo and Video Standard Set [34] consists of videos from 50 patients and the University of California, San Diego (UCSD) Facial Nerve Database consists of videos from 15 patients clinically diagnosed with facial palsy. Thus, a total of 65 individuals with facial palsy was used in this study. Additionally, videos of 50 healthy controls were gathered at Stanford University. In this section, we provide a brief description of the three databases used, collection of healthy control data, experimental setup, and facial cue tasks. Table I summarizes the data used in this study.

**TABLE 1.** Summary of all study participants (*n*=115).

| Category | eFACE | Palsy Type | No. of Subjects | Database |
| --- | --- | --- | --- | --- |
| Healthy Controls | 96 – 100 | None | 10<br>34<br>6 | MEEI<br>Stanford<br>UCSD |
| Palsy | < 96 | Flaccid<br>Synkinetic | 11<br>4 | UCSD |
| Near-Healthy Palsy | 91 – 95 | Flaccid<br>Synkinetic | 5<br>5 | MEEI |
| Moderate Palsy | 80 – 90 | Flaccid<br>Synkinetic | 5<br>5 | MEEI |
| Severe Palsy | 70 – 79 | Flaccid<br>Synkinetic | 5<br>5 | MEEI |
| Complete Palsy | < 60 | Flaccid<br>Synkinetic | 5<br>5 | MEEI |

#### MEEI Facial Palsy Photo and Video Standard Set

This open-source dataset, hosted on the Sir Charles Bell Society website, is a standardized set of facial photographs and videos that capture the spectrum of flaccid and nonflaccid, or synkinetic, facial palsy [34]. Standard intake protocol was followed, including a clinician assessment of facial function (eFACE) as well as a set of photographs and video. Exclusion criteria included prior facial reanimation surgery or extensive facial scarring, currently active chemodenervation, and bilateral facial palsy. Cases from both flaccid and nonflaccid (aberrantly regenerated or synkinetic) states were included resulting in 25 flaccid palsy patients and 25 synkinetic palsy patients.

Participants who consented to enroll in the standard set were categorized by their eFACE score into the following: healthy, near-healthy, mild, moderate, severe and complete flaccid or nonflaccid facial palsy. The degree of palsy was quantified using eFACE, House-Brackmann (HB), and Sunnybrook scales by two expert clinicians in the field. For this study, all 50 individuals with facial palsy were used and the remaining 10 individuals were used as healthy controls.

#### UCSD Facial Nerve Database

IRB approval was obtained from the Office of IRB Administration at University of California San Diego (UCSD) (protocol 210007, originally approved 05-07-2021 and has been approved on annual review since). For this database, patients who have facial palsy greater than a HB score of I, or an eFACE score of *<* 86, are currently being recruited from the UCSD Facial Nerve Clinic. A total of 15 individuals with known facial palsy were used in this study. Of these 15 patients, 11 have flaccid palsy and 4 have synkinetic palsy.

#### Healthy Controls

A total of 50 healthy controls were used in this study. Ten healthy controls (5 male and 5 female) that were included in the MEEI Facial Palsy Photo and Video Standard Set were used. The remaining 40 individuals (20 male and 20 female) recruited consists of healthy volunteers, who have no facial movement disorders. IRB approval was obtained from the Office of IRB Administration at Stanford (protocol 74656, approved 04-08-2024) to recruit these healthy controls.

#### The Facial Nerve Exam

All the participants included in this study performed the same standard facial nerve (FN) exam, in which the subject was requested to perform the following facial cue for a few seconds before returning back to face at rest.

1. Face at rest: used as baseline
2. Raise their eyebrows
3. Gently close their eyes
4. Forcefully close their eyes
5. Gentle smile
6. Full effort smile
7. Pucker their lip
8. Show their bottom teeth

Fig. 1a presents the intended facial muscle activation for each of the given cues.

**Fig. 1.**
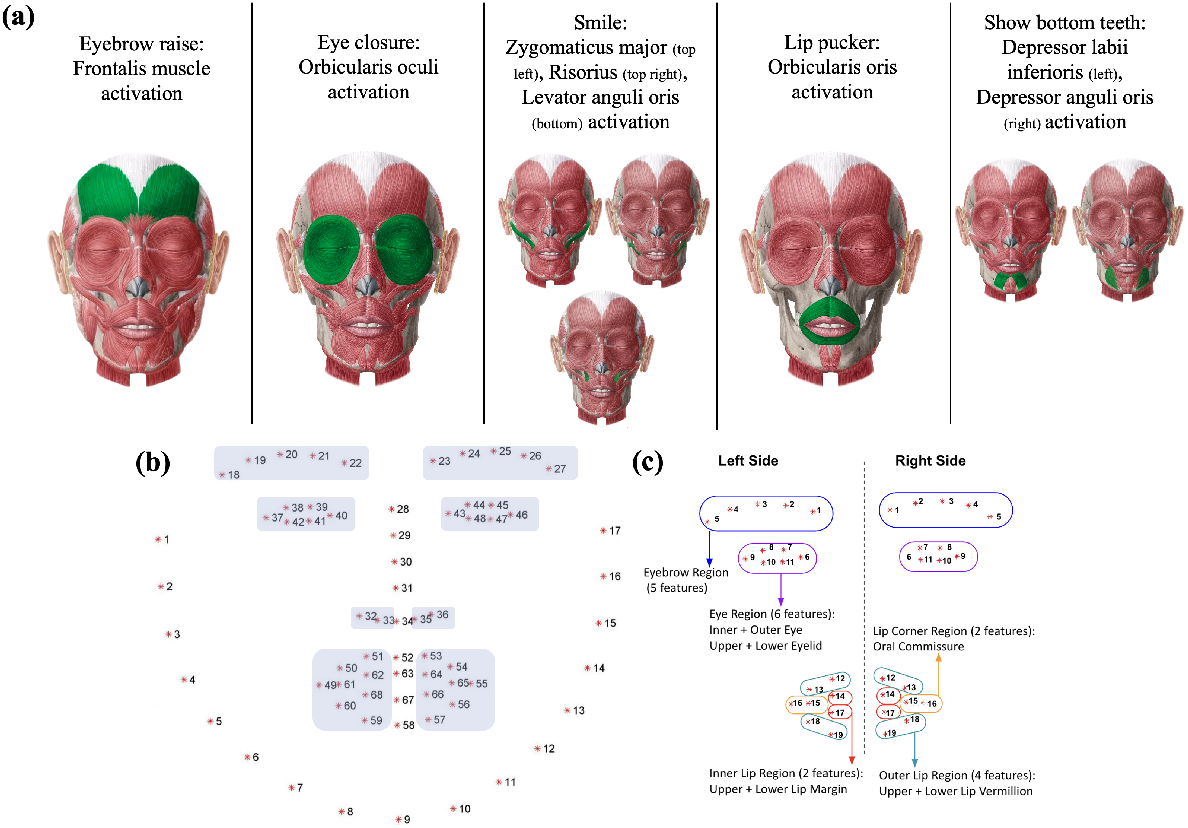
(a) The five facial cues given during the facial nerve (FN) exam and their intended facial muscle activation, where the region of interest is highlighted in dark green. (b) The pre-trained facial landmark detector inside the *dlib* library is used to estimate the location of 68 (x, y)-coordinates that map to facial structures on the face. The highlighted points refer to the 38 features used for the computational measures. Diagram of the 38 facial features that were extracted from each side of the patient’s face during the recorded FN exam.

### B. Data Processing

#### Facial Landmarks Extraction

Face detection was performed on each frame using histogram of oriented gradients (HOG) + a support vector machine (SVM), and a total of 68 facial landmarks were extracted by using the open-source software Dlib [35]. While there are newer and more modern face detection models that exist, such as Haar-based techniques and multi-task cascaded convolutional neural networks (MTCNN), Dlib (which uses HOG + SVM) has shown to outperform Haar and MTCNN when it comes to processing speed and lowlight conditions [36]. To reduce the influence of interfering information, such as background, brightness and head posture, gray-scale normalization and geometric normalization methods were used to normalize feature information from each frame [37].

#### Gray-Scale Normalization and Feature Extraction

1. Extract the number of frames in the input video.
2. For each frame in the video:
  a. Convert the color input image to gray scale.
  b. Detect the face on the image using the open-source and publicly available *dlib* and *OpenCV* Python libraries.
  c. Store the coordinates in the selected frame.
  d. Save coordinates for all the frames in the video as a .csv file for future data processing.

#### Geometric Normalization

1. The x and y coordinates of each facial landmark was subtracted from the average coordinates during the face at rest cue.
2. The x and y coordinates of every facial landmark was then subtracted from the midpoint of the subject’s face as identified by *dlib*. This midpoint corresponds to the (x,y) coordinate of landmark 34 from Fig. 1b.
3. These positional differences were normalized by subtracting their mean values over the first 100 frames, ensuring consistent alignment across patients.

The pre-trained facial landmark detector inside the *dlib* library is used to estimate the location of 68 (x, y)-coordinates that map to facial structures on the face. Note that while the *dlib* shape predictor is trained to detect 68 points (Fig. 1b), only 38 of them were used in this work to reduce redundancy and focus on dynamic features. These 38 points were reorganized, as seen in Fig. 1c, to proceed with the computational measures. *Segmenting Data for Specific Cues*: The start and end times of each FN exam cue in a subject’s video were manually annotated. Instead of trimming the videos to capture only the facial expressions, the entire video was retained, and all frames within the annotated time windows for each cue were used for cue-specific analyses. These annotated time points were then converted in specific frame numbers based on the frames per second rate of the input video. Using these frame numbers, the corresponding landmark coordinates were segmented and stored as a Python dataframe for further processing.

### C. Implementing supervised learning for hypothesis testing Feature selection

To probe aspects of facial symmetry, we extracted the x and y positional coordinates of 38 facial landmarks and computed their differences relative to the midline of the face. Specifically, for each landmark, we calculated the difference in x and y coordinates between the left and right sides of the face. These positional differences were normalized by subtracting their mean values over the first 100 frames, ensuring consistent alignment across patients. Thus, for each participant, we obtain a matrix *<u>Y</u>* ∈ ℝ^*n*×*d*^, where *n* = 38, representing the number of landmarks, and *d* corresponds to the number of frames in the patient video. We observe statistically significant differences between the patients and healthy controls (*p <* 0.0001, corrected for multiple comparisons) in some key facial regions, such as the lip corner (oral commissure) and outer and inner lip features, between the average y-position difference, as shown in Fig. 2a.

**Fig. 2.**
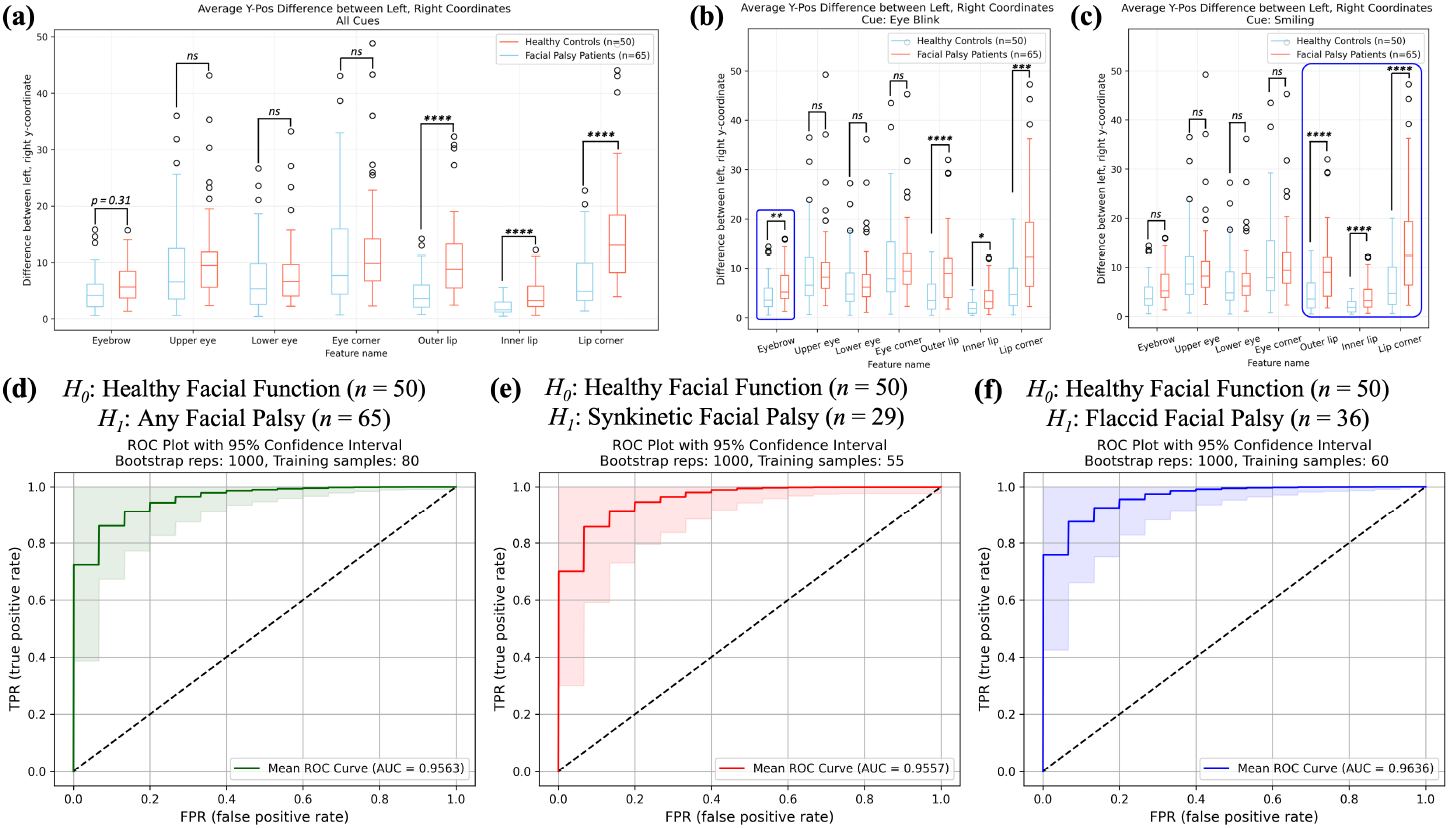
(a) Average difference of y-coordinates on the left and right side of the face for all the facial palsy patients and healthy controls over the entire course of the FN exam. (b) Average difference of y-coordinates on the left and right side of the face for all facial palsy patients and healthy controls during the blinking cue. (c) Average difference of y-coordinates on the left and right side of the face for all facial palsy patients and healthy controls during the smiling cue. All significance values were corrected for multiple comparisons (*: *p <* 0.05, **: *p <* 0.01, ***: *p <* 0.001, ****: *p <* 0.0001,). d-f presents ROC curves of 1000 bootstrap samples for the log-likelihood ratio test when using the Wasserstein barycenter approach between (d) facial palsy patients (*n=65*) and healthy controls (*n=50*), where the mean ROC AUC was 0.955, (e) synkinetic patients (*n=29*) and healthy controls, where the mean ROC AUC was 0.9557, and (f) flaccid patients (*n=36*) and healthy controls, the mean ROC AUC was 0.9637.

A similar process can be applied to the landmark coordinate data segmented at selected cues, resulting in 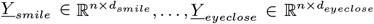. During each cue, we observe statistically significant differences in varying facial regions, with highest significance in context-specific regions of interest. For example, during the blinking cue, there is a significant difference for the eyebrow features (*p <* 0.05, corrected for multiple comparisons) and lip features (*p <* 0.001, corrected for multiple comparisons) (Fig. 2b). Whereas, during the smile cue, there is a significant difference for all the lip features (*p <* 0.0001, corrected for multiple comparisons) (Fig. 2c).

#### Setup for binary classification

Our setup for binary classification can be described as having labeled data (*<u>Y</u>* ^(1)^, …, *<u>Y</u>* ^(*k*)^), which are our arbitrary feature vectors for every subject in ℝ^*k*^, and (*H*_1_, …, *H*_*k*_) such that *H*_*i*_ ∈ {0, 1}, where 0 is our label for healthy facial function and 1 for facial palsy. The goal of binary classification is to find a decision function that allows us to use the input features *<u>Y</u>* ^(*i*)^ that provide the corresponding class labels *H*_*i*_. This can be viewed as a hypothesis testing problem, where it is assumed that under *H*_0_, *<u>Y</u>* has a joint density *f*_*<u>Y</u>*_ (*<u>y</u>*; *θ*_0_ : *<u>µ</u>*_0_, Σ_0_), and under *H*_1_, *<u>Y</u>* has a joint density *f*_*<u>Y</u>* (*<u>y</u>*_; *θ*_1_ : *<u>µ</u>*, Σ_1_).

Both hypothesis classes were modeled as a multivariate Gaussian distribution, with mean *<u>µ</u>* and covariance matrix Σ, namely *<u>Y</u>* ~ 𝒩 (*<u>µ</u>*, Σ). Then, note that its density is given by:

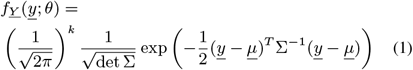

We can utilize supervised learning to partition the samples for each class, where we use 70% of the dataset for training and the remaining 30% for testing. The training dataset has the labels for their associated classes and so we can calculate distribution parameters, *θ*_*i*_, for every training sample under *H*_0_ and under *H*_1_.

The receiver operating characteristic (ROC) curve offers one way to measure the effectiveness of prediction by calculating the area under the curve (AUC), which can be interpreted as the probability that the result of the test of a randomly chosen individual with facial palsy is more indicative of paralysis than that of a randomly chosen healthy control [38]. To determine the reliability and performance of the classification, bootstrapping (*n = 1000)* was used to provide a range of ROC curves, and thus calculate an average AUC score, helping us understand how robust the model evaluation is to variations in the dataset.

### 2) Clustering probability distributions using Wasserstein barycenters

To perform model fitting to provide one distribution for any class, we employ an approach that acknowledges the distinct statistical pattern in each human subject within that class. This approach is robust to subject-to-subject intraclass feature variability.

Optimal transport theory is a geometrically meaningful way of measuring distances between probability distributions, and it has recently become important in applications in data science and machine learning [39]. In this work, we employ the concept of barycenters to find the “center of mass” of a collection of probability measures, where distances between each are determined by the optimal transport Wasserstein distance.

Consider a set *P*_1_, *P*_*k*_ of elements, with their associated weights *λ*_1_, …, *λ*_*k*_, satisfying *λ*_*i*_ *>* 0 and 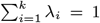, belonging to a metric space *M*. The barycenter *P* ^*^ on this metric space (*M, d*_*M*_) is defined as [40]:

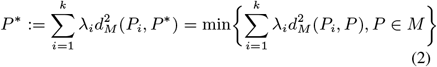

If we were to consider the set *P*_1_, …, *P*_*k*_ of elements to be probability distributions, where the metric space (*M, d*_*M*_) corresponds to the 2-Wasserstein distance, then *P* ^*^ becomes the Wasserstein barycenter [41]. When *P*_1_, …, *P*_*k*_ are all multivariate Gaussian distributions with parameters (*µ*_*i*_, Σ_*i*_), then the mean of the Wasserstein barycenter is simply 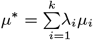 and the covariance Σ^*^ is the only positive definite matrix Σ satisfying the equation [42]:

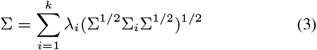

Using the probability distributions of each training sample in either hypothesis class, the Wasserstein barycenter was calculated to define distribution parameters, 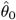 and 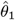, for each distribution, where 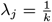. Then, with the test dataset, where the label of the subject was unknown, a likelihood ratio test was performed on each sample, using the learned parameters, 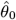 and 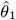 from the training dataset. A likelihood ratio test was performed under the two Gaussian Wasserstein barycenters:

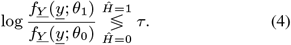

This supervised learning method for hypothesis testing was used to classify between: 1) healthy controls and all facial palsy patients, 2) healthy controls and synkinetic facial palsy patients, and 3) healthy controls and flaccid facial palsy patients.

### D. Identifying regions of dynamic facial asymmetry using specific facial cues

#### 1) Pearson correlation coefficient to define facial symmetry

Pearson correlation coefficients are well-suited for capturing linear relationships between facial landmarks, quantifying the degree of linear dependence between them. These coefficients provide an effective means of assessing proportional changes in facial features during dynamic expressions. The magnitude of the Pearson coefficient reflects the strength of the correlation, offering a quantitative measure of how facial features move in tandem.

From the 38 facial landmarks described in Fig. 1b, Pearson correlation coefficients were calculated for each of the corresponding right and left landmark, resulting in a coefficient vector, *<u>v</u>* ∈ ℝ^19^. These coefficient vectors were computed either:

1. Across the Entire FN Exam: To assess overall facial symmetry during the full duration of the exam, a single set of coefficients was calculated over all frames in the video, capturing global facial movement patterns.
2. For Specific Facial Cues: For analyses targeting individual expressions (e.g., eyebrow raise, smile), the coefficients were calculated within the time windows corresponding to the given cues. The start and end frames for each cue were manually annotated, and only the frames within those intervals were used to compute the coefficients.

This dual approach allows for both comprehensive and cue-specific evaluations of facial symmetry, providing insights into overall facial function, as well as nuanced movements associated with specific expressions. Furthermore, heatmaps were created to spatially localize levels of synchrony for the 19 pairs of facial landmarks across the face for each of the five cues executed.

#### 2) Mahalanobis distance as a metric to determine context specific asymmetry

To compute how much a patient deviates from healthy facial function, the normalized Mahalanobis distance (MD) was calculated for each FN exam cue. This metric identifies which of the cues has the most significant dynamic asymmetry when compared to healthy function and gives insight into the specific region and context, or facial cue, of asymmetry.

Though seven cues were provided in the FN exam, the two eye closure cues and the two smile cues were combined to reduce redundancy. Therefore, the five facial cues of interest include eyebrow raise, eye closure, smile, lip pucker, and showing lower teeth. For each facial cue, the MD was used to measure the degree of error from healthy facial function based on the 19 Pearson coefficients of the symmetric facial landmarks. The MD takes into account the variability and correlation structure of the data, making it suitable for multivariate analysis. The following is how MD was computed for each cue:

1. The Pearson correlation coefficient vectors was calculated for all 50 healthy controls, *<u>V</u>* ∈ ℝ^19×50^
2. The mean of the Pearson coefficients for all healthy controls was calculated, *<u>µ</u>* ∈ ℝ^19^. This represents the average correlation values across all facial regions.
3. The covariance matrix of the Pearson coefficients for all healthy controls was determined, Σ ∈ ℝ^19×19^. This describes the relationships and variability among the different facial regions.
4. For the patient of interest, the MD was calculated at each of the 5 cues: 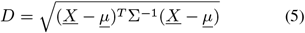

where *<u>X</u>* is the vector of Pearson coefficients for the patient of interest, *<u>µ</u>* is the mean vector of all the healthy controls, and Σ is the covariance matrix of all the healthy controls.
5. Once all the distances were calculated for every cue 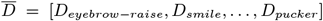, the five distances were normalized allowing us to determine which cue resulted in maximum deviation from healthy facial function.

The MD provides a measure of how far away the facial palsy patient’s data is from the average healthy controls’ function by considering the multivariate distribution of healthy controls’ Pearson coefficients. A higher MD indicates that the facial palsy patient’s function is more divergent from the healthy control range.

### E. Using multivariate outlier detection to identify facial palsy

#### 1) Wasserstein barycenter distance as metric to define facial palsy

The Wasserstein distance quantifies the minimum “work” required to transform one probability distribution into another. As it defines a metric space within probability measures, it can be applied to compare the distribution of facial feature positions during expressions in individuals with varying degrees of facial palsy to a reference distribution representing healthy facial function. To calculate the Wasserstein distance of one multivariate Gaussian distribution to another Gaussian, *d* := *W*_2_(𝒩 (*<u>µ</u>*_1_, Σ_1_); *𝒩* (*<u>µ</u>*_2_, Σ_2_)), the following closed-form equation can be used [39, eq 2.41]:

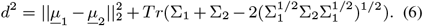

To extend the notion of the MD that was used for the *<u>v</u>* ∈ ℝ^19^ of Pearson correlation coefficients to the patient specific y-position coordinates, Wasserstein distances were calculated. This distance to the cue-specific barycenter of all healthy controls was calculated for each subject in this study (*n* = 115):

1. The cue in which a subject deviated most from the average healthy function was identified by using normalized MDs. The cue with the largest MD was identified as having maximum deviation.
2. For this cue, the Wasserstein barycenter that summarizes the Gaussian distribution for all healthy controls was calculated. See (2), (3).
3. The Wasserstein distance between the individual subject’s Gaussian distribution at the cue and the Wasserstein barycenter for all healthy controls was calculated.

This distance serves as a representative measure of the dissimilarity between an individual’s facial movements and the typical facial expressions of the healthy population. We hypothesized that patients with facial palsy would have noticeably larger Wasserstein distances from the averaged healthy controls barycenter at that cue. As such, the Wasserstein barycenter distance can be useful for outlier detection because it provides a measure of how far a particular distribution is from the central tendency represented by the barycenter. To account for outliers in the healthy controls, Wasserstein distances that were above two standard deviations from the mean were not used for the following linear regression models. As such, seven healthy controls were removed for the model selections to follow.

To determine the effectiveness of using Wasserstein distances to distinguish between healthy controls and facial palsy patients, binary classification via a threshold-based approach was performed using a train-test split of 80% −20%. Bootstrapping (n = *1000*) was used to provide a range of ROC curves, and the mean AUC was used to evaluate the classification performance.

#### 2) Linear regression to predict clinical scores of facial palsy

To further assess the clinical utility of facial landmarks, we aligned our analysis with validated scores widely used in the field of Otolaryngology–Head and Neck Surgery: the House-Brackmann (HB) scale, the Sunnybrook Facial Grading System, and the eFACE scale [43], [44].

- *House-Brackmann (HB) Score*: The HB scale, a six-point system approved as the reference standard for grading facial palsy, ranges from grade I (healthy facial function) to grade VI (complete paralysis).
- *Sunnybrook Facial Grading System*: This composite scale (0–100) evaluates facial symmetry at rest, voluntary movement, and synkinesis, with higher scores indicating better facial function.
- *eFACE*: The eFACE generates a continuous score (0–100) based on the degree of asymmetry and movement during facial expressions, with higher scores reflecting near-healthy facial function.

Some patients in the databases had all three scores annotated by clinical experts in the field, while all patients had HB scores available. This allowed us to evaluate how well our computational methods correlate with established clinical metrics and to ensure the clinical relevance of our findings. The healthy controls were assigned a HB score of I and a 100 on the Sunnybrook and eFACE scales since they exhibited healthy facial function in all areas. As such, the calculated Wasserstein distances were used as the independent variable and the clinical scores are the dependent variable. Wasserstein distances were calculated for all the participants (*n*= 108 after 7 healthy control outliers removed).

### F. Application to Surgical Cases

To demonstrate how our proposed machine learning algorithms can be applied to track FN recovery over time and inform clinical decisions, we analyzed two surgical cases: an iatrogenic FN injury and FN weakness due to a cancerous tumor. The cases presented here highlight the utility of dynamic facial function analysis in both early detection of recovery and in guiding surgical decisions.

#### 1) Case 1: Tracking of subtle onset facial nerve recovery for an iatrogenic patient

Iatrogenic FN Injury has a reported incidence of 11% to 40% [45], where rates can vary depending on the type of surgical procedure done and expertise of the surgeon. There is consensus among peripheral nerve surgeons that the critical window following acute injury during which nerve repair is feasible is up to 72 hours. There are cases where this timeline may be extended several weeks with contemporary instruments [46]. However, subtle changes in FN performance are difficult to evaluate both from an observational manner and from current measurements, such as scoring scales like HB or EMG, which only yields 33% accuracy in traumatic injury patients [47].

This case presents an iatrogenic patient (IP), Fig. 5, who underwent a tumor excision where the surgeon was not certain whether a cranial nerve was damaged or not (stretch injuries can mimic transection or more severe damage). Videos of IP performing the FN exam were collected the week following injury and then again 3.5 weeks after.

#### 2) Case 2: Tracking facial nerve function pre- and post-facial nerve sacrifice and repair of a head and neck cancer patient

Subtle, progressive and non-resolving facial palsy can be the sole initial clinical manifestation of perineural spread of head and neck cancer occurring along the facial nerve (CN VII), and be misdiagnosed as a benign process such as Bell’s Palsy in cancer survivors, particularly in the absence of a discrete tumor. Facial paralysis continues to pose a significant concern during cancer resection and facial nerve interventions are often not prioritized amidst urgent oncologic treatments [48]. Detection of subtle changes in facial nerve function preoperatively could greatly aid surgical planning as the optimal time for facial nerve reconstruction is at the time of tumor resection.

This case presents a patient with a right parotid malignant tumor (CP), Fig. 6, who underwent a radical right parotidectomy with a FN sacrifice, lateral temporal bone resection, and neck dissection to remove the tumor. Videos of CP performing the FN exam were taking 3 weeks prior to surgery when they started developing FN weakness, 8 months after surgery to remove the tumor and repair FNs, and 2 years after the surgery.

## III. Results

### A. Performance of binary classification of facial palsy

The average AUC, with a 95% confidence interval, was calculated for the three binary classification scenarios. Fig. 2d-f presents the ROC curves along with the average AUC values for each scenario when using the Wasserstein barycenter approach to cluster the probability distributions for each patient. The following AUC value was calculated for each classification scenario: Fig. 2d) healthy controls and facial palsy patients: 0.9563, Fig. 2e) healthy controls and synkinetic facial palsy patients: 0.9557 and Fig. 2f) healthy controls and flaccid facial palsy patients: 0.9636.

### B. Patient-specific assessment of dynamic facial asymmetry is more robust during specific contexts

The Pearson coefficients were calculated at the five facial cues of interest for all 50 healthy controls, and the average values of all 19 coefficients that correspond to the y-position of the landmarks in Fig. 1b are presented in Table II. It is observed that all 19 features reveal high positive correlations between the right and left side y-coordinates during the duration of the FN exam. The average Pearson coefficient heatmaps for all healthy controls during the five cues are presented in Fig. 3a, where dark blue corresponds to a positive correlation of 1 and dark red corresponds to a negative correlation of −1. During all the cues, there are strong positive correlations, above 0.90 for each cue’s regions of interest:

**TABLE 2.**
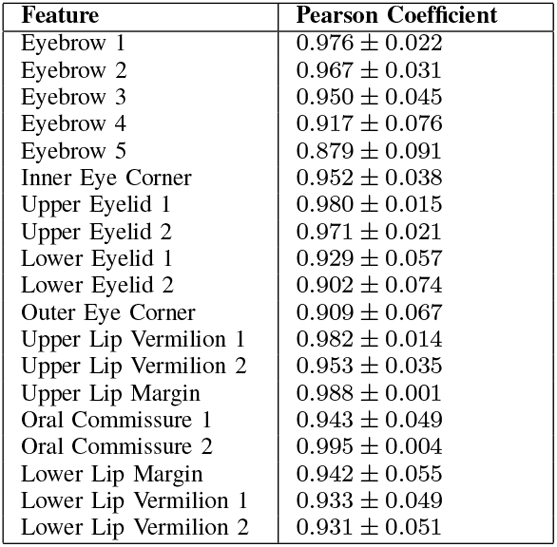
Average of 19 y-position pearson correlation coefficients for healthy controls (N=50).

**Fig. 3.**
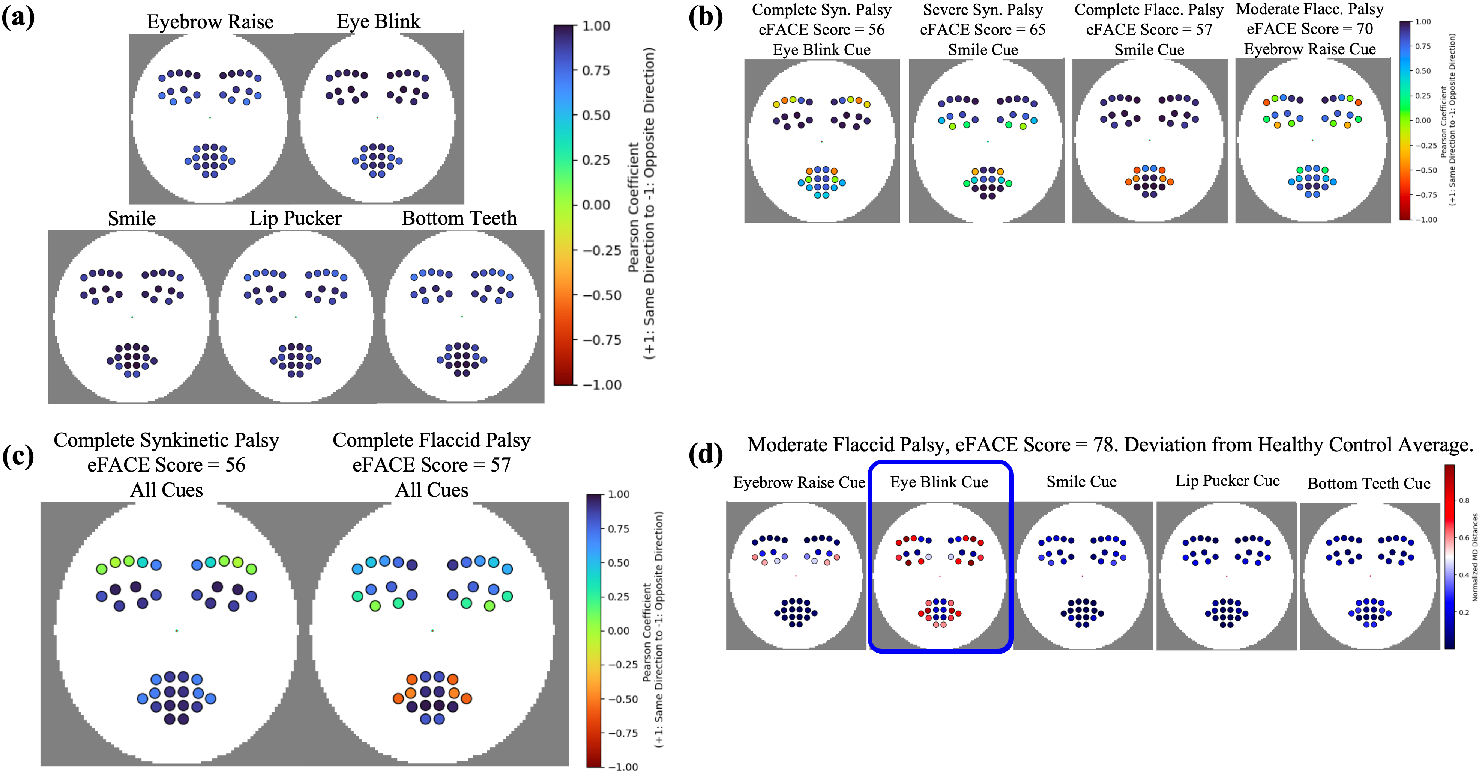
(a) The average and standard deviation of the 19 Pearson correlation coefficients for all healthy controls used in this study (*n=50*). These are the correlation coefficients calculated for the entire course of the FN exam. (b) Plotting Pearson coefficients for the 19 features during specific cues for each patient. These heatmaps represent individual patient dynamic asymmetry when comparing the changes in y-coordinates of the left side and right side of their face. In the order of left to right presents a complete synkinetic palsy patient (SP1) (eFACE = 56), severe synkinetic palsy patient (eFACE = 65), complete flaccid palsy patient (FP1) (eFACE = 57), and moderate flaccid palsy patient (eFACE = 70). (c) Plotting Pearson coefficients for the 19 features during the whole time series for SP1 and FP1. (d) Using normalized Mahalanobis distances (MDs) to compute how much a patient deviates from healthy facial function during each cue. The metric identifies the eye blink cue to deviate the most from healthy facial function for this patient with moderate flaccid palsy (FP2) (eFACE score = 78).

1. *r*_*y*−*eyebrow*_ = 0.911 ± 0.052 during the eyebrow cue
2. *r*_*y*−*eyeclosure*_ = 0.952± 0.022 during the eye closure cue
3. *r*_*y*−*lipregion*_ = 0.934 ± 0.056 during the smile cue
4. *r*_*y*−*lipregion*_ = 0.900 ± 0.043 during the lip pucker cue
5. *r*_*y*−*lipregion*_ = 0.911± 0.066 during the show bottom teeth cue

Fig. 3b presents four patients from the MEEI Facial Palsy Photo and Video Standard Set with varying types of facial palsy and their corresponding heatmaps during specific FN exam cues (please view Section V regarding images of patients in figures). The complete synkinetic palsy patient (SP1) (left in Fig. 3b) presents palsy in the eyebrow region and upper lip vermilion and oral commissure, and this is captured in both the magnitude and direction of the Pearson coefficients in these regions: *r*_*y*−*eyebrow*_ = −0.280 ± 0.187, *r*_*y*−*uppervermilion*_ = − 0.228 ± 0.327, and *r*_*y*−*oralcommissure*_ = 0.55. Similarly, the complete flaccid palsy patient (FP1) (second from right in Fig. 3b) presents palsy in the oral commissure and upper lip vermilion, and this is captured in the Pearson coefficients of these regions: *r*_*y*−*oralcommissure*_ = −0.54 and *r*_*y*−*uppervermilion*_ = −0.525 ± 0.009.

When comparing the above results to the Pearson coefficients of the whole time series, rather than specific cues, we lose some of the spatial specificity and sensitivity of asymmetry but the algorithm can still identify key regions of facial palsy. Fig. 3c presents the heatmaps of the same two patients described above but for the full duration of the FN exam. Over the whole time course, SP1 has the following Pearson coefficients for the initially identified regions of asymmetry: *r*_*y*−*eyebrow*_ = 0.032 ± 0.0488, *r*_*y*−*uppervermilion*_ = 0.685 ± 0.007, and *r*_*y*−*oralcommissure*_ = 0.67. While FP1 has the following Pearson coefficients for the initially identified regions of asymmetry: *r*_*y*−*oralcommissure*_ = −0.58 and *r*_*y*−*uppervermilion*_ = −0.515 ± 0.077.

In the case of FP1 the regions of asymmetry are apparent during the duration of the whole FN exam, but as evident in SP1, there are specific contexts or facial cues in which their irregular motor movement is most evident. To better determine under which context a patient’s palsy is most noticeable in, the Mahalanobis distance (MD) was used to measure the degree of error or how off a patient is from healthy facial function. Fig. 3d presents a patient with moderate flaccid palsy (FP2) performing all the five facial cues, with the normalized MD values for each 19 symmetric landmarks spatially plotted on top. It can be observed that in some cues, such as the smile and lip pucker cue, FP2’s palsy is not as evident as it is during the eye closure and eyebrow raise cue. A higher MD indicates that landmarks at that specific cue are more divergent from the healthy range, allowing us to identify at which context patients differ most from the healthy controls. In FP2’s case, the highest MD is attributed to the eye closure cue, indicating that this is when the patient’s palsy deviates most from healthy function (as defined in Fig. 3a).

By creating separate heatmaps for each facial cue during the FN exam, clinicians can focus on specific regions of interest. This feature-specific analysis helps identify asymmetries or patterns related to particular facial landmarks, aiding in the assessment of facial expressions or cues.

### C. Performance of using Wasserstein distances to predict clinical scores

The mean AUC from the n=1000 iterations was 0.9129 (Fig. 4a). A linear least-squares regression model was fit between a patient’s Wasserstein distance and their given 1) HB score (*n* = 108; *r* = 0.611; *p <* 0.0001), 2) eFACE score (*n* = 93; *r* = − 0.656; *p <* 0.0001), and 3) Sunnybrook score (*n* = 93; *r* = − 0.621; *p <* 0.0001). Fig. 4b-d present the scatter plots of the Wasserstein distance estimating the three different facial grading scales commonly used by clinicians, with their respective linear least-squares regression and corresponding 95% confidence interval.

**Fig. 4.**
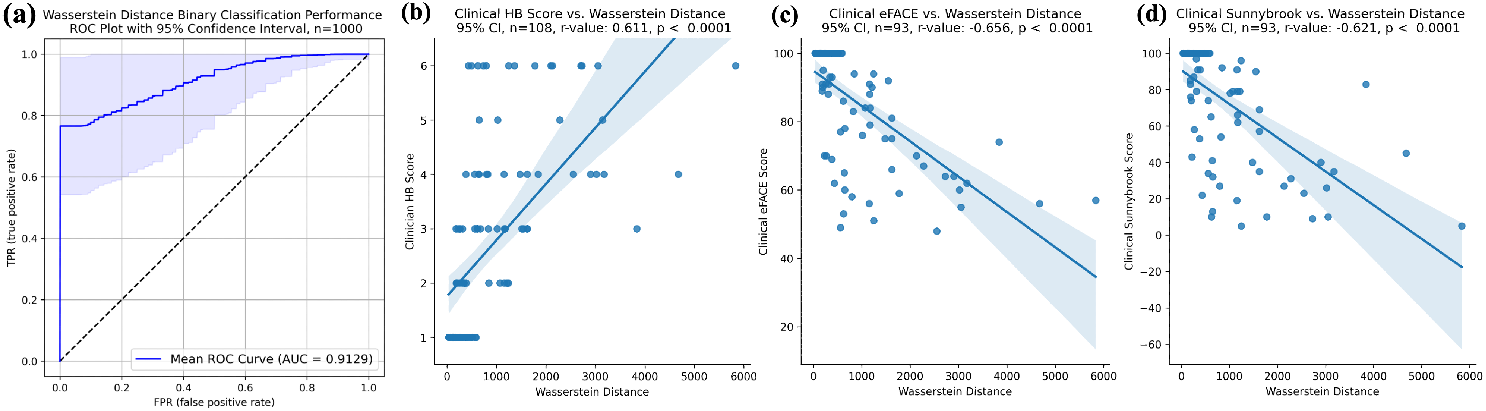
(a) Presents the ROC curve to test the accuracy of using Wasserstein distances to classify between facial palsy patients and healthy controls (n = 1000 iterations). The mean AUC of the ROC curve was 0.9129. Scatter plots between the Wasserstein distance and the given clinician (b) HB score (*n* = 108; *r* = 0.611; *p <* 0.0001), (c) eFACE score (*n* = 93; *r* = *−* 0.656; *p <* 0.0001), and (d) Sunnybrook score (*n* = 93; *r* = *−* 0.621; *p <* 0.0001). The blue line is a linear least-squares regression and the light blue sharing is the corresponding 95% confidence interval.

### D. Application to Surgical Patient Cases

#### 1) Case 1: Tracking of subtle onset facial nerve recovery for an iatrogenic patient

Table III presents the Pearson correlation coefficients during both trials, and the corresponding heatmaps are presented in Fig. 5a for the 0 week time point and Fig. 5d for 3.5 week time point (please view Section V regarding images of patients in figures). Differences in Pearson coefficient values are observed specifically during the smile cue and the lip pucker cue; the coefficient magnitudes of the eyebrow and eye features increase after 3.5 weeks and are statistically significant (*p <* 0.05) (Table III), indicating increased facial dynamic symmetry. However, since the magnitudes of the coefficients for the features of the lip region, specifically the lower lip margin, decreased significantly (*p <* 0.05), IP has not reached full healthy facial function.

**Fig. 5.**
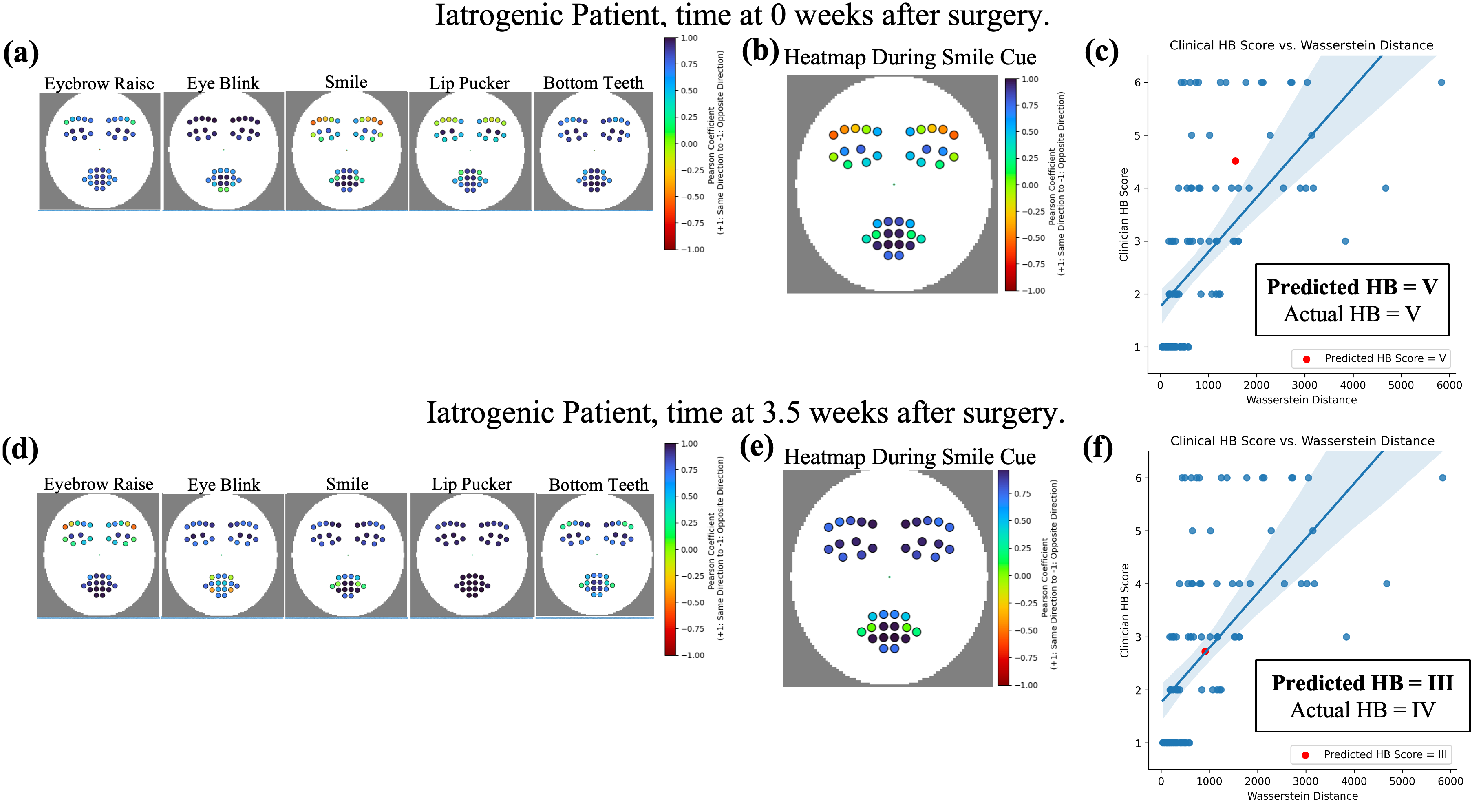
(a-c): Analysis for the iatrogenic patient (IP) at 0 weeks post-injury. a: Plotting Pearson coefficients for the 19 features during specific cues. b: Tracking Pearson coefficients during the cue that exhibited maximum deviation from healthy function: smile. c: Using the linear regression model to predict IP’s HB score, which was predicted to be V. (d-f): Analysis for the IP at 3.5 weeks post-injury. d: Plotting Pearson coefficients for the 19 features during specific cues. e: Tracking Pearson coefficients during the same cue exhibited maximum deviation from healthy function as in b: smile. Increases in Pearson coefficient magnitudes are observed over time, indicating FN recovery. f: Using the linear regression model to predict IP’s HB score, which was predicted to be III. IP’s predicted score drops from an initial HB V to a HB III, and these scores align with the clinician given score with an MSE of 0.5

**TABLE 3.** Pearson coefficients at the regions of interest during the smile cue for iatrogenic patient.

| Feature | Coefficient at 0 weeks | Coefficient at 3.5 weeks |
| --- | --- | --- |
| Eyebrow 1 | 0.554 | 0.973 |
| Eyebrow 2 | -0.035 | 0.937 |
| Eyebrow 3 | -0.285 | 0.804 |
| Eyebrow 4 | -0.441 | 0.715 |
| Eyebrow 5 | -0.547 | 0.744 |
| Upper Eyelid 1 | 0.497 | 0.944 |
| Upper Eyelid 2 | 0.771 | 0.946 |
| Outer Eye Corner | -0.049 | 0.815 |
| Oral Commissure 1 | 0.757 | 0.741 |
| Lower Lip Margin | 0.359 | 0.209 |

$p \leq 0.05$ (\*) for all Pearson coefficients.

The MD was used to identify the smile cue as the context under which palsy was most apparent for IP. With this context, the Wasserstein distances were calculated for IP and the linear least-squares regression model was used to predict HB scores. The model predicts IP to have a HB score of V at week 0 (Fig. 5c) and a HB score of III at week 3.5 (Fig. 5f). These predicted scores align with the clinician assigned scores, which were HB V at week 0 and HB IV at week 3.5, with a mean square error (MSE) of 0.5 for the two predictions.

#### 2) Case 2: Tracking facial nerve function pre- and post-facial nerve sacrifice and repair of a head and neck cancer patient

Fig. 6 tracks CP’s initial onset of FN weakness and FN function 8 months and 2 years after surgery (please view Section V regarding images of patients in figures). The initial FN weakness was initially most apparent under the eye closure cue, where slight weakness is observed in the lower eye regions and oral commissure areas (Fig. 6a). Fig. 6b, c present worsening paralysis in the eyebrow region after surgery with the maximum deviation contexts varying from the smile cue at 8 months to the bottom teeth cue at 2 years. With the maximum deviation contexts determined by the MD algorithm, CP’s Wasserstein distances were calculated at each time point. The linear least-squares regression model was used to predict HB scores both before and after surgery. The model predicts an initial HB score of III when they first develop FN weakness 3 weeks prior to surgery (Fig. 6d). The model predicts a HB score of IV 8 months post-surgery, when CP developed mild synkinesis (6e). The predicted score increases to a VI 2 years after surgery (Fig. 6f). These predictions align with the clinician assigned scores, which were HB III 3 weeks post-surgery, HB IV 8 months post-surgery, and HB III 2 years post-surgery, with a MSE of 3.0 for the three predictions.

**Fig. 6.**
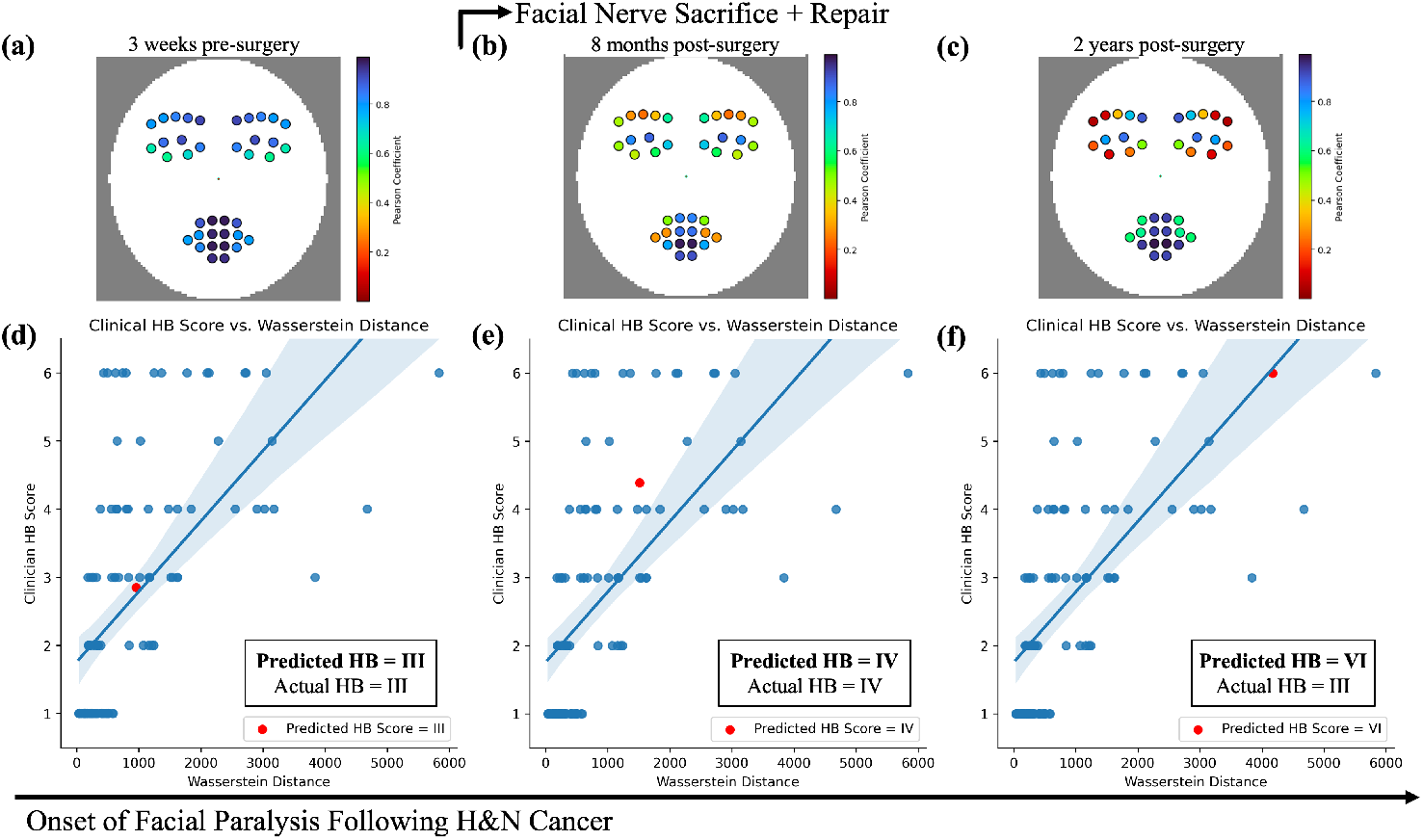
Tracking CP’s FN recovery, along with the Pearson coefficient heatmaps, over the course of when they developed FN weakness (a) 3 weeks prior to surgery (b) 8 months after their tumor removal and facial reanimation surgery (c) and 2 years after surgery. Using the linear regression model to predict CP’s HB score over the course of their clinic visits. (d) The model predicts an initially HB score of III when they first develop FN weakness 3 weeks prior to surgery. (e) The model predicts a HB score of IV 8 months post-surgery, when CP developed mild synkinesis. (f) The model predicts a HB score of VI 2 years post-surgery, when CP developed moderate synkinesis. These three predictions align with the clinician given score with an MSE of 3.0.

## IV. Discussion

Current methods of facial nerve (FN) assessment rely heavily on clinician experience and subjective scales, such as the House-Brackmann (HB) or Sunnybrook scales, often applied to static photographs [49], [50]. These approaches may miss subtle dynamic changes, such as interval asymmetry or segmental weakness, and are prone to inter-observer variability, which has been reported to reach 64% for severe facial palsy cases (HB III or worse) [51]. Although machine learning (ML) algorithms for facial analysis have advanced significantly [18], their application to static images of facial paralysis patients often results in significant landmark inaccuracies, limiting their clinical utility [20], [21]. Addressing these challenges, recent studies have shown that dynamic video analysis reduces landmark errors compared to static image analysis [32], [37].

In this study, we report for the first time a novel technique of using Wasserstein barycenters and Mahalanobis distances to develop ML techniques to classify between healthy controls and facial palsy patients in videos, along with accurate metrics to predict commonly used clinical grading scales like the HB and Sunnybrook. By clustering probability distributions using barycenters, we are utilizing an approach that is robust to outliers [52], [53]. This is important to account for while using human subject data, where inter-class variability and outliers are inevitable. Furthermore, the barycenter technique accounts for the full range of facial landmarks and their probabilities over time, providing a richer and more discriminative representation of facial function. This probabilistic approach makes our technique more robust for distinguishing between facial palsy subtypes and assessing severity, offering a more nuanced understanding of dynamic facial movements compared to traditional methods.

We also demonstrate that Pearson correlation coefficients can be used to quantify facial symmetry over the duration of videos. By analyzing spatio-temporal landmark data over specific cues, these coefficients provide a robust statistical measure of the linear relationship between facial movements, allowing us to identify regions of asymmetry. This video-based machine learning analysis could, for the first time, characterize the dynamic properties of various facial movements, such as smiling versus raising eyebrows, without requiring manual assessment, thereby significantly advancing our understanding of facial function. This cue-specific analysis of facial function was crucial in determining the Wasserstein distances that helped develop prediction models to map back to clinical scores. This cue-specific analysis not only improves model accuracy but also highlights critical regions of dysfunction, offering clinicians valuable insights to optimize diagnostic and surgical strategies.

Finally, we present surgical case studies to demonstrate the practical utility of our proposed algorithms in clinical settings. In Case 1, our algorithm successfully detected subtle improvements in facial nerve recovery for an iatrogenic patient. The visualization metrics helped track recovery over the 3.5-week period, proving valuable for decisions on whether reanimation surgery was necessary. In Case 2, involving a cancer patient, the algorithm captured the progression of facial nerve weakness both before and after surgery, with the regression model aligning fairly well with clinician-assigned HB scores. This case underscores the potential of our approach to track and predict facial nerve function over time, providing clinicians with objective insights to guide decisions regarding tumor resection and post-surgery management. These cases highlight the real-time clinical applicability of our algorithms in assessing facial nerve function and improving decision-making for various patient scenarios.

### A. Limitations

There are limitations to this study. Landmark errors persist in videos of facial palsy patients, particularly around challenging features such as eyelids during blinking, facial rhytids, and facial hair. Additionally, patient effort can affect movement range, especially in facial palsy patients, who may consciously or subconsciously avoid large movements, such as smiling, to mask their asymmetry. Other errors may arise from unstable camera setups or other noise artifacts. Improved preprocessing steps after landmark extraction will enhance accuracy in future iterations of this work.

While manual annotation was used in this study, the methodology provides a framework for future development of an automatic approach to detect the start and end of facial expressions. Such automation could leverage current computer vision techniques to recognize facial movements or specific cues in untrimmed videos, eliminating the need for manual intervention and improving usability in home settings.

While the model’s predictions for the iatrogenic patient yielded an MSE of 0.5, it produced an MSE of 3.0 for the cancer patient. This discrepancy partly arises from inherent limitations in the House-Brackmann (HB) scale itself. In particular, the HB scale does not fully capture the range of synkinetic palsy severity once a patient moves beyond mild synkinesis, rated at Grade III. If a patient presents with severe synkinesis, the scale essentially caps this at HB Grade IV, leading to a mismatch between the predicted and clinically assigned scores. This highlights the need for clinician expertise in cases where synkinesis is prominent; a refined approach that incorporates more granular clinical observations could better accommodate these nuances. Moreover, uncertainty analysis, as explored in other clinical contexts [54], could further improve our methodology by flagging ambiguous data for clinician review. This approach recognizes the inherent variability in clinical assessments and aligns with the “art of medicine,” where gray areas often require expert judgment. Future work may focus on integrating uncertainty metrics and potentially extending the HB scale to account for advanced synkinesis, ensuring that the model reflects the full spectrum of facial nerve recovery while enhancing both its reliability and clinical applicability.

### B. Future Directions

Expanding the database of healthy controls and facial palsy patients in a controlled setting will help capture the full spectrum of dynamic facial function and refine prediction models. Discerning the distinctive patterns in specific types of palsy can allow for more accurate prediction models, advancing personalized treatment strategies and improving outcomes for individuals affected by facial paralysis.

It is important to note that none of the existing facial grading systems discussed in this study possess predictive capabilities for patient outcomes. This is a significant limitation in the field, as these systems primarily assess the current state of FN function without forecasting recovery or deterioration. A promising future direction of our research is to develop a predictive model based on the video-based analysis we propose.

By leveraging dynamic facial function data, we could predict potential trajectories of patient outcomes, enabling clinicians to make more informed decisions about interventions and to monitor recovery progress more accurately over time.

Dynamic facial analysis has promising future applications, especially with recent advances in telemedicine. Secure, patient-driven, HIPAA-compliant iOS telemedicine apps now leverage computer vision and AI for various uses, such as tracking knee arthroplasty rehabilitation [55], monitoring epilepsy [56], and assessing liver steatosis [57]. With over half a million head and neck cancer survivors in the US, this algorithm could enable patients to track their facial function post-therapy using a telemedicine tool powered by machine learning. This tool could reduce diagnostic delays, provide objective data for clinicians, and empower patients across all socioeconomic backgrounds, given the widespread availability of smartphones. By advancing the evaluation of facial nerve function from static, subjective assessments to dynamic, objective analyses, this work represents a meaningful step toward more precise, patient-centered care in facial paralysis.

## V. Conclusion

This study introduces a dynamic and quantitative framework for video-based analysis of facial function, leveraging advanced machine learning techniques such as likelihood ratio tests, optimal transport theory, and Mahalanobis distances. These methods enable precise classification of facial palsy types, identification of asymmetric regions, and assessment of severity in facial paralysis compared to healthy controls. Our results demonstrate that video analysis provides a significantly more accurate and detailed assessment of facial movements than previously reported, along with the ability to identify specific facial cues where asymmetry or paralysis is most evident. Pearson correlation coefficients, and visualizing them as spatial heatmaps, could serve as a clinical tool to quantify facial synchrony and monitor subtle changes in facial nerve function over time. Additionally, the novel application of Wasserstein barycenters offers an objective approach to map facial asymmetry severity directly to clinical scores, improving the ability to assess FN recovery and surgical outcomes longitudinally.

The clinical relevance of this work lies in its potential to transform facial paralysis management. By providing objective, quantitative data, the proposed methods can enhance surgical decision-making, improve the timing and outcomes of reanimation surgeries, and ultimately improve the quality of life for patients. Furthermore, this approach facilitates early detection of FN recovery or deterioration, supporting timely interventions and better long-term outcomes. This study sets the stage for integrating precision-based video analysis into clinical practice, providing a scalable, automated tool to reduce reliance on subjective scoring and manual annotation. These advances mark a significant step forward in facial nerve assessment and offer a foundation for future innovations in patient-centered care for facial paralysis.

## VI. Addendum

Due to pre-print screening requirements, images of the facial palsy subjects discussed in this study will only be released in the final publication. Readers may contact the corresponding author prior to final publication to request access to these materials.

## Data Availability

All data produced in the present study are available upon reasonable request to the authors.

## Notes

### Competing Interest Statement

The authors have declared no competing interest.

### Funding Statement

This study was funded by Stanford Graduate Fellowship in Science & Engineering.

### Author Declarations

IRB of University of California, San Diego gave ethical approval for the photo and video-based assessment at the facial nerve center (protocol #210007, originally approved 05-07-2021, UCSD Facial Palsy Database). Office of IRB Administration at Stanford gave ethical approval for the recruitment and video collection of healthy controls (protocol #74656, approved 04-08-2024).

### Summary of Updates

We have made substantial revisions to enhance clarity, methodology, and structure. We clarified our video processing methodology, stating that videos were segmented but not trimmed, ensuring dynamic motion data was preserved. Additionally, we specified that Pearson correlation coefficients were computed both globally and for specific cues. The manuscript was further restructured by clearly separating Methods from Results, introducing case studies in advance, and organizing the Discussion into distinct subsection, including Limitations and Future Directions. We removed the nested regression model due to its limited clinical utility and confirmed that p-values were Bonferroni corrected, now explicitly noted in figure captions. Lastly, we updated language for inclusivity, reformatted figures and tables for clarity, and aligned content with preprint standards. These revisions strengthen the manuscript's rigor and applicability.

